# Effective Stimulation Sites and Networks for Substantia Nigra Pars Reticulata Deep Brain Stimulation for Auditory Verbal Hallucinations in Schizophrenia

**DOI:** 10.1101/2025.04.09.25325419

**Authors:** Min Jae Kim, Ankur Butala, Yousef Salimpour, Akira Sawa, Martijn Figee, Ki Sueng Choi, David Schretlen, Kelly A. Mills, Nicola Cascella

## Abstract

Deep brain stimulation (DBS) of the substantia nigra pars reticulata (SNpr) is under investigation for managing auditory-verbal hallucinations (AVH) in treatment-resistant schizophrenia (TR-SZ). We assessed acute AVH suppression during initial SNpr-DBS programming in three TR-SZ patients and mapped associated brain network engagement using normative connectomes. One-month post-implantation, monopolar stimulation at each electrode contact was evaluated for its effect on AVH severity. Volumes of tissue activation (VTA) were integrated with normative structural and functional connectivity data to generate individualized network maps. Among 86 VTAs, stimulation sites associated with greatest AVH relief localized to left anterior-dorsal and right posterior-ventral SNpr. Greater AVH suppression correlated with structural connectivity to sensorimotor cortex, precuneus, angular and supramarginal gyri, and functional connectivity to the mediodorsal thalamus, orbitofrontal cortex, anterior cingulate, and dorsolateral prefrontal cortex. These preliminary results highlight specific SNpr subregions and circuits linked to acute symptom reduction, supporting the potential of network-targeted DBS for TR-SZ.

## Introduction

Schizophrenia is a complex neuropsychiatric disorder with global prevalence of 0.28%, causing significant deterioration in quality of life and significantly reduced life expectancy ^1^. About 60% to 80% of patients with schizophrenia experience auditory and verbal hallucinations (AVH) ^2^. While antipsychotic drugs are considered first-line therapies, 25-30% of patients with AVH remain refractory to pharmacotherapy^3^, resulting in an unmet need for alternative interventions for persons with treatment resistant schizophrenia (TR-SZ).

For the last decade, neuromodulation therapies such as Deep Brain Stimulation (DBS) have shown great promise for many neuropsychiatric disorders, including obsessive-compulsive disorder (OCD) ^4–7^ and major depressive disorder (MDD) ^8–10^, among other novel indications. Ongoing investigations aimed at improving DBS in these neuropsychiatric disorders have focused on identifying precise and accurate stimulation targets ^8,11,12^, as well as elucidating key neural circuitries associated with the disease pathology that are involved in effective DBS. These studies have provided a mechanistic explanation on how DBS intervenes in a larger neural circuitry to achieve clinical benefit and allowed optimization of its effectiveness in future studies through iterative refined targeting of specific structures and/or brain networks.

In contrast to OCD and MDD, neuromodulatory applications in TR-SZ have been limited. DBS for TR-SZ has been attempted, via anatomical targeting of anterior cingulate cortex, nucleus accumbens, and habenula; albeit with variable outcomes within and between targets limited by small sample sizes. We have recently shown substantial improvement in AVH in a patient with TR-SZ following DBS of the substantia nigra pars reticulata (SNpr) ^13^. SNpr was chosen as a target for AVH because we hypothesized that high-frequency stimulation of GABAergic inhibitory SNpr projections to the dorsomedial thalamus may normalize activity in connected cortical regions involved in AVH generation, such as the superior temporal gyrus. Here, we explore the network mechanism of SNpr DBS for AVH in three patients with TR-SZ using normative functional and structural connectomes to investigate which SNpr connections are involved in AVH suppression during acute stimulation.

## Results

### Demographics and Clinical Information

Demographic and clinical information of three patients is listed below in **Supplementary Material 1**. All patients experienced auditory hallucinations with a primary diagnosis of schizophrenia.

During the monopolar review, patient 1 had total 27 unique stimulation parameters tested (14 left side, 13 right side, 0.5 – 2 V, 60 μsec, 130 Hz). For patient 2, 41 unique stimulation parameters were tested (24 left side, 17 right side, 0.5 – 4.9 mA, 60 μsec, 130 Hz). Finally for patient 3, 18 unique stimulation parameters were tested (9 left side, 9 right side, 0.9 – 2.5mA, 60 μsec, 125 Hz). This resulted in a total of 86 VTAs (47 left, 39 right).

After all the VTAs and paired AVH Likert scale scores were pooled together, AVH scores underwent Z-score transformation to allow group-level analysis. The raw Likert AVH ratings and Z-scored AVH ratings were highly correlated (r=0.635, p < 0.0001) shown in **Figure 1A**, confirming that the transformation could reliably capture the variance of AVH scores across different stimulation parameters.

**Figure 1.**
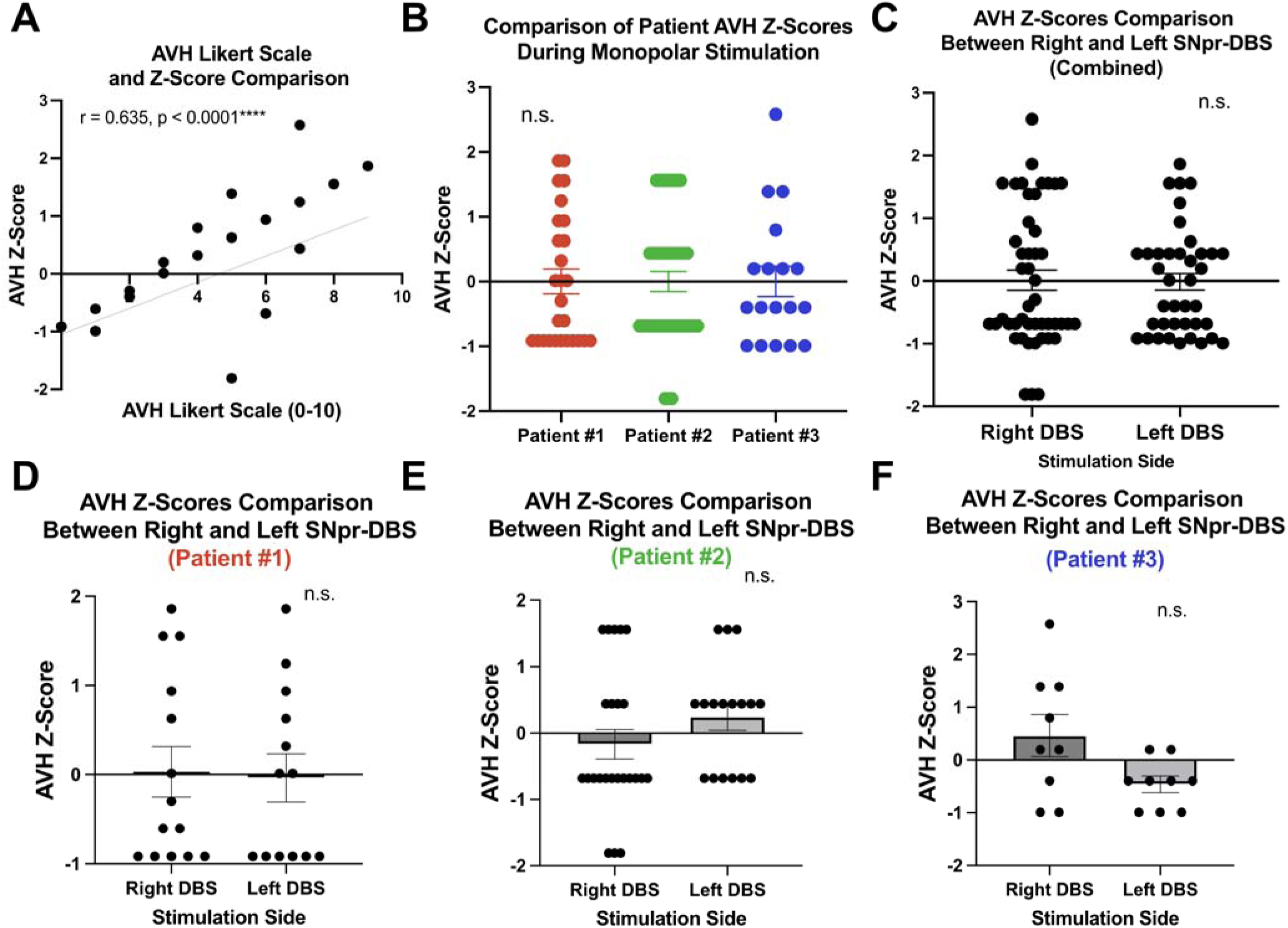
Comparison of AVH Scores from DBS. **(A)** Correlation between raw Likert-Scale AVH scores and AVH Z-scores. **(B)** Comparison of AVH Z-scores across the patients. Comparison of AVH Z-scores across hemisphere of stimulation across all patients in the cohort **(C**) and within individual patients **(D-F).**

AVH Z-scores were also compared between the three patients to explore if variance was driven by a certain patient. One-way ANOVA test, followed by post-hoc Tukey’s test for pairwise comparison, revealed non-significant differences of scores between patients (F = 0.1739 (2,83), p=0.84) shown in **Figure 1B**. Further, the distribution of AVH scores between each stimulated hemispheres were not statistically different across patients (**Figure 1C**), or within each patient (**Figure 1D-F**).

### Mapping AVH Scores Across SNpr Stimulation Sites

Reconstructed anatomical electrode locations are visualized in **Figure 2** relative to atlas-defined SNpr ^14^. VTA models weighted by z-scored AVH scores were used to create MEI for each hemisphere (**Figure 3**). For left-side stimulation, lower than average AVH scores (i.e MEI regions > 0) were seen when the anterior-dorsal portion of SNpr was stimulated, and higher-than-average AVH scores (i.e MEI regions < 0) were associated with stimulation at the posterior-ventral border of SNpr. However, for right-side stimulation the posterior-ventral SNpr was associated with optimal AVH suppression (**Figure 3B – C)**. Centroid coordinates of “sourspot” and “sweetspot” of MEI are shown in **Figure 3D**.

**Figure 2.**
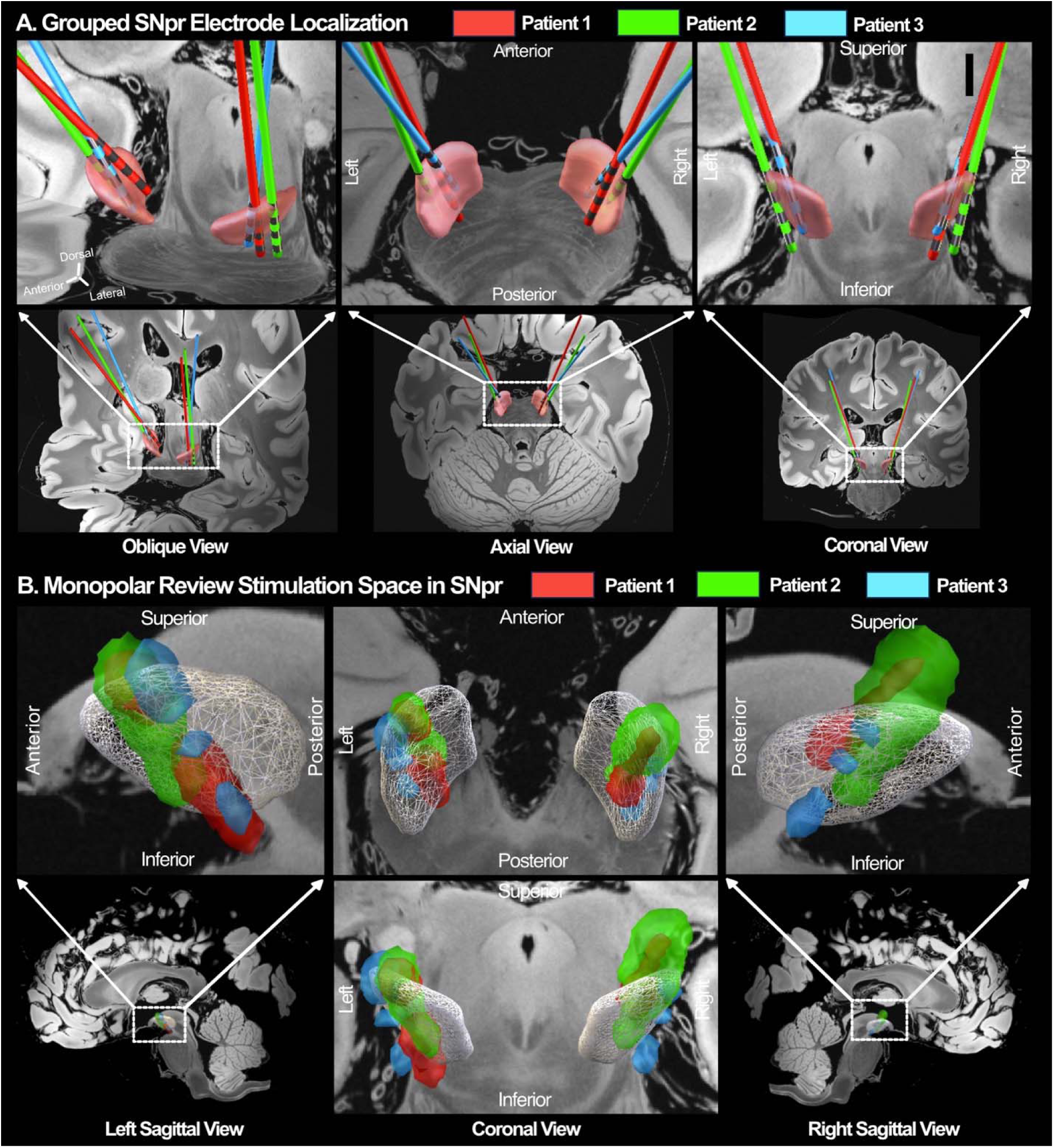
DBS Electrode Localization and Monopolar Stimulation Space. **(A)** DBS electrodes of three patients are localized and visualized relative to SNpr atlas (Pauli et al, 2018) (23) with 7T 100 *u*mm *ex-vivo* MRI backdrop (Edlow et al, 2019). **(B**) Stimulation Space in SNpr. During acute monopolar review of each patient, each stimulation parameters was used to model VTA. Stimulation space was defined as area captured by all VTAs produced per patient in both hemispheres shown relative to SNpr (white mesh).

**Figure 3.**
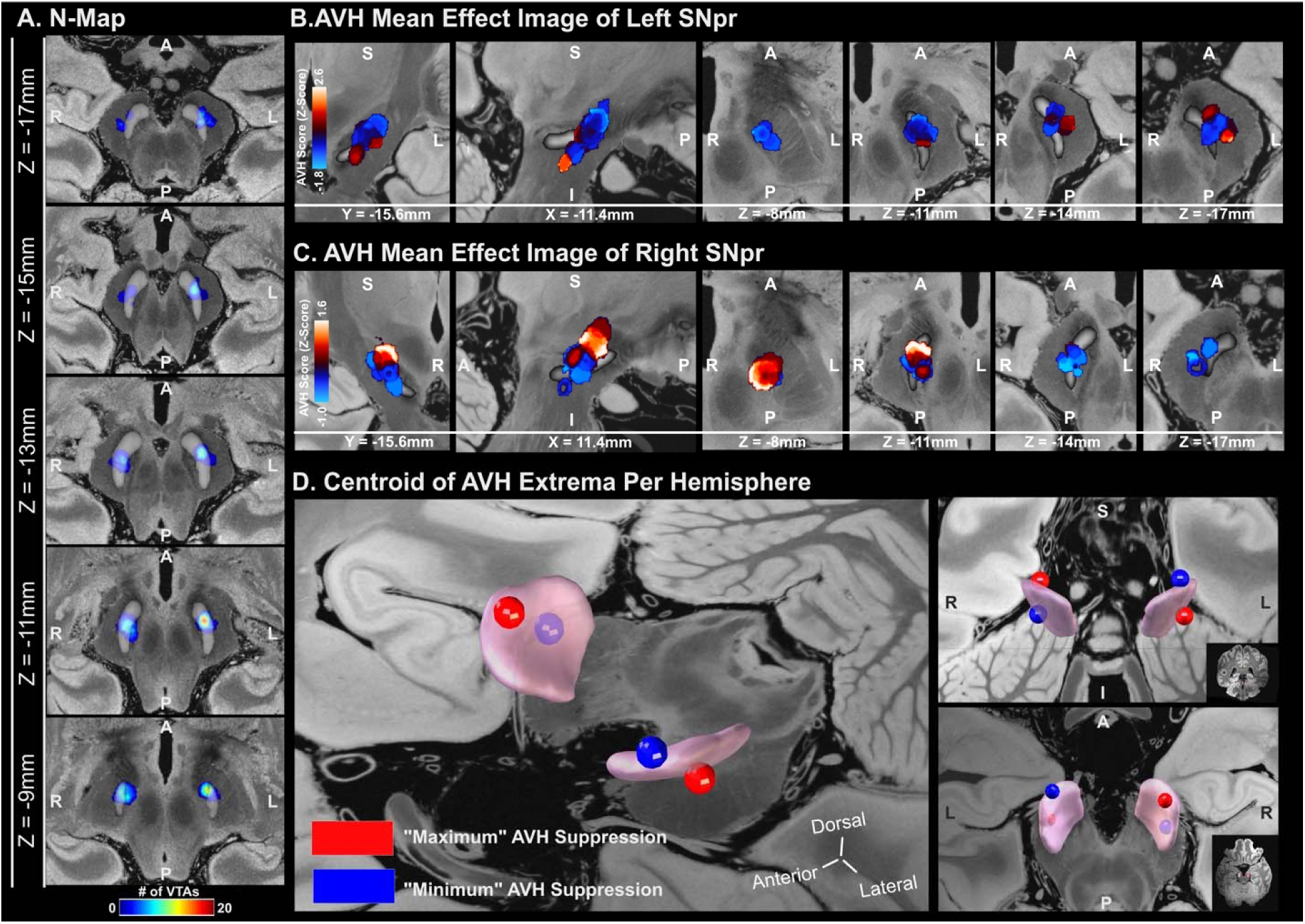
AVH Mean Effect Image (MEI) and VTA Density Map. **(A)** Voxel-wise density of VTAs is represented as N-Map bilaterally. Mean AVH z-scores attributed to VTAs occupying each voxel are shown as MEI for **(B) left** and **(C) right SNpr**. **(D)** Centroid of MEI where top (red sphere) and bottom 5% extremes (blue sphere) of voxel intensities are located are mapped relative to SNpr. R= Right, L = Left, S = Superior, I = Inferior.

**Figure 4.**
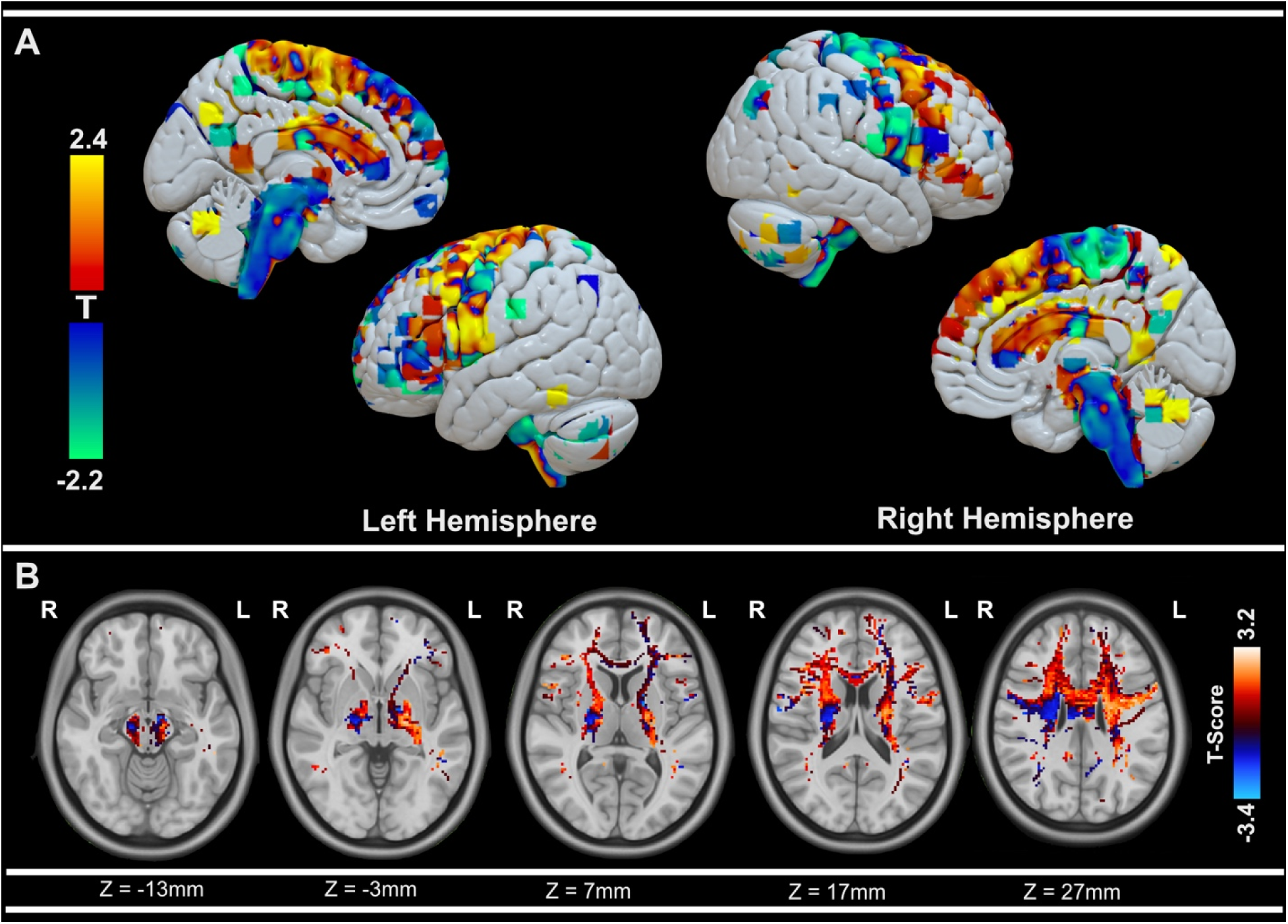
AVH Structural Connectivity T-Map. **(A)** Cortical rendering of T-Map of whole brain voxel-wise structural connectivity streamlines numbers relative to bilateral VTAs associated with more or less AVH suppression. **(B)** T-score distributions across subcortical regions at different axial planes. Positive T-values (red – yellow) are areas associated with more t AVH suppression, and negative T-values (blue - green) as areas with less AVH suppression. R= Right, L = Left.

For the right-sided MEI, the centroid coordinate of “sourspot” was located at X=11.15mm, Y=-13.97, Z=-9.28mm encompassing regions with mean z-score of 1.32, and “sweetspot” at X=11.46mm, Y=-18.98mm, Z=-14.58 of regions with mean z-score of −0.95 (**Figure 3D**). For the left-sided MEI, the centroid coordinate of “sourspot” was located at X=-12.38mm, Y=-17.58, Z=-14.84mm encompassing regions with mean z-score of 1.13, and the “sweetspot” was located at X=-11.70mm, Y=-12.33mm, Z=-8.96 of regions with mean z-score of −1.43.

### Structural Connectivity Associated with Acute AVH Suppression

When seeding VTAs from both hemispheres, lower AVH scores (FDR-corrected p-value = 0.05) were significantly correlated to increased streamlines from VTAs reaching left sensorimotor cortex (t=-3.21,p=0.0019), left angular gyrus (t=-2.66,p=0.0094), left precuneus (t=-2.06,p=0.042), and right supramarginal gyrus (−2.85,p=0.0056).

Less AVH suppression was observed when there was increased connectivity of VTAs to right sensorimotor cortex (t=2.81,p=0.006), supplementary motor area (t=2.16, 0.03), rolandic operculum (r = 2.15,p=0.03), and temporal regions including the right hippocampus (t=2.64,p=0.010), superior temporal gyrus (t=3.32,p=0.0013), and middle temporal gyrus (t=2.18,p=0.032)

### Streamline Fibertracks Associated with AVH Suppression

To better characterize how distribution of streamline fibers corresponds to AVH suppression, fiberfiltering analysis was adopted. Distribution of fibers predictive of AVH scores were further explored separately for right and left SNpr stimulation. For right SNpr stimulation, only fibers assigned with top and bottom 25% of T-scores were considered. Amongst the total number fibers (516) passing through the right SNpr VTAs, the majority of fibers (98.8% (510)) were occupied by fibers with negative T-scores (−2.26 ± 0.0039) transversing posterior-lateral portion of SNpr.

Right sided fibers associated with higher AVH severity (6.17 ± 0.13) occupied a 1.16% of total number of distributed fibers with, transversing the anterior-medial portion of the right SNpr (**Figure 5A**). Similar to the analysis with pooled VTAs above, mean T-scores of fibers were significantly correlated with AVH score (r=0.75,p=0.0002) and scores could be predicted during leave-one-out (r=0.32,p=0.041) and 5-fold (r=0.56,p=0.0004) cross-validation, shown in **Figure 5B**.

**Figure 5.**
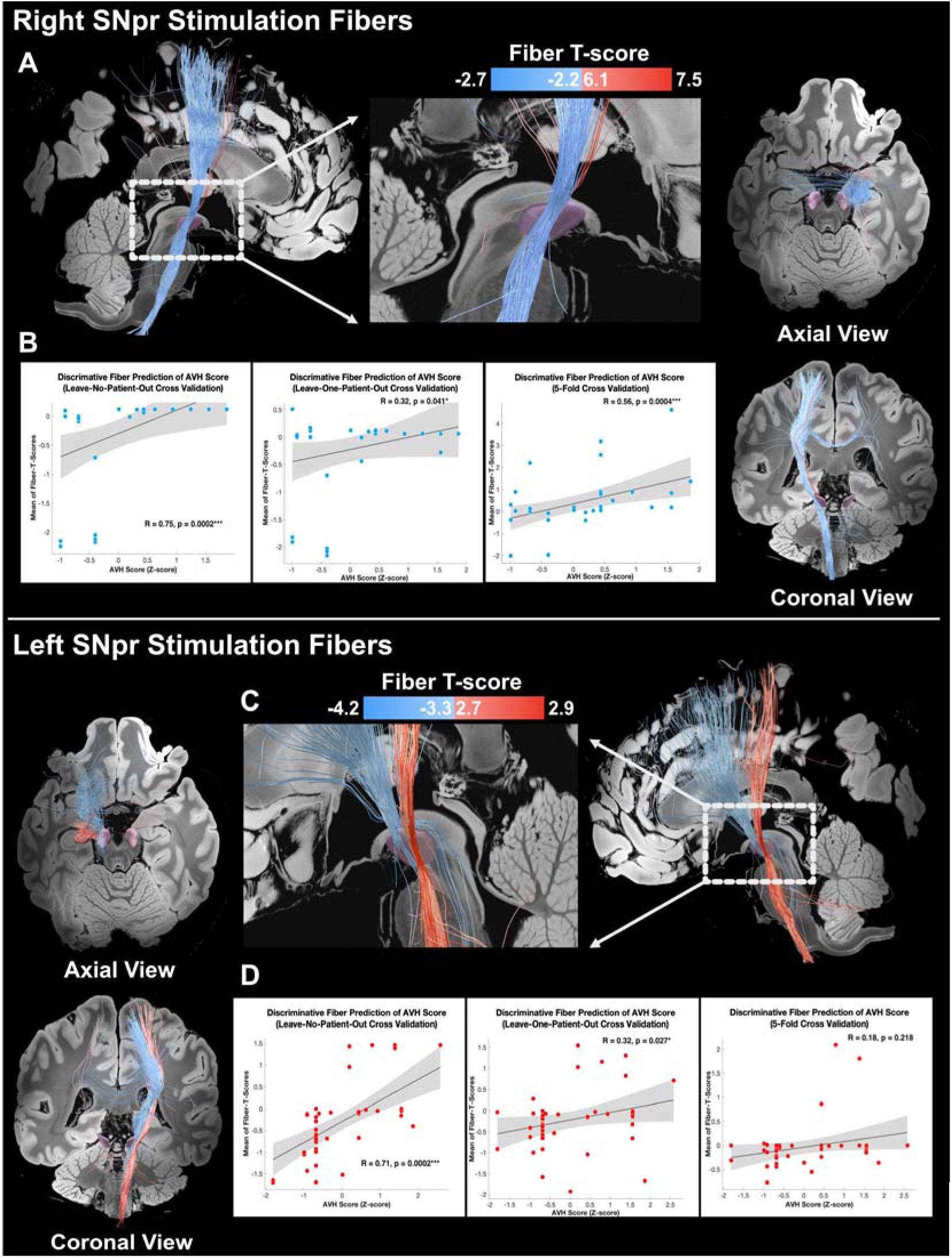
Streamline Fibers Associated with AVH Severity (blue = more suppression, red = less suppression) from Unilateral VTAs. Streamlines separately seeded from VTAs produced from **(A)** right-sided and **(C)** left-side stimulation. Mean T-scores attributed to each fiber were used to predict AVH scores paired with each VTAs. The performance of the prediction model was tested in naïve condition, leave-one-out, and 5-fold cross-validation scheme for separately for (**B)** and **(D)** left stimulation. R=Right. L= Left.

For left side stimulation, amongst the 740 total fibers (fibers with top and bottom 25% of t-scores) passing through left SNpr, 64% (472) of fibers assigned negative scores (−3.38 ± 0.011) associated with lower AVH scores transversed through the anterior-medial portion of SNpr (**Figure 5C**). On the other hand, posterior SNpr was occupied by fibers assigned positive t-scores (2.72 ±0.011) occupying 36% of total fibers. Mean T-scores of the fibers were also strongly correlated with AVH scores (r=0.71,p=0.0002), and this model performed significantly well during leave-one-out (r=0.32,p=0.027) but not during 5-fold (r=0.18,p=0.22) cross-validation shown in **Figure 5D**.

### Functional Connectivity Associated with AVH Suppression

Similar to T-maps generated based on structural connectivity using seed-based streamline counts, T-maps were produced using functional connectivity (FC) measures to further survey potential polysynaptic connections not explained from SC profiles (**Figure 6**). Positive voxels in T-maps represent regions were increased in FC relative to VTAs is correlated with higher AVH scores (i.e. less suppression of AVH), and negative voxels with lower AVH scores (i.e. more AVH suppression).

**Figure 6.**
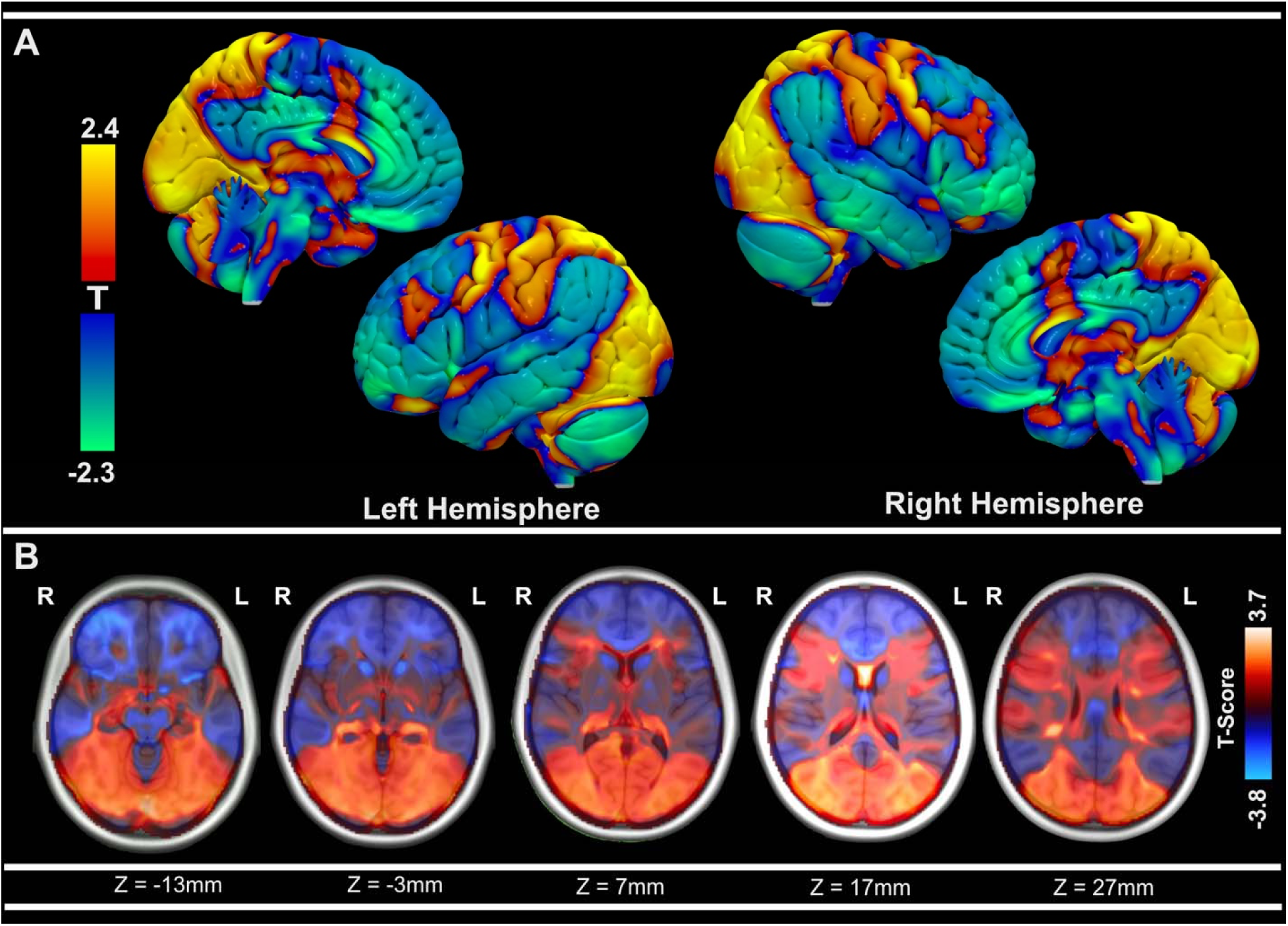
AVH Functional Connectivity T-Map. **(A)** Cortical rendering of T-Map of whole brain voxel-wise functional connectivity strength relative to bilateral VTAs associated with AVH severity. **(B)** T-score distributions across subcortical regions at different axial planes. Positive T-values (red – yellow) are areas associated with less AVH suppression, and negative T-values (blue - green) as areas with more AVH suppression. R= Right, L = Left.

When seeding VTAs from both hemispheres, increased FC to the following cortical regions was associated with lower AVH scores (FDR corrected p-value = 0.05): bilateral anterior orbitofrontal cortex (OFC) (right: t=-3.02,p=0.0033, left: t=-2.14,p=0.03), superior anterior cingulate cortex (ACC) (right: t=-2.84,p=0.0058, left: t=-2.86,p=0.0053), middle cingulate cortex (right: t=-2.24,p=0.028, left: t=-2.64,p=0.0099), right middle frontal gyrus (t= −2.11,p=0.038), and right inferior frontal gyrus pars orbitalis (t=-2.07,p=0.04). In addition to cortical engagement, increased connectivity to thalamic regions, the lateral pulvinar nucleus (right: t=-2.41,p=0.018, left: t=-3.17,p=0.002), right inferior medial dorsal nucleus (t=-2.05,p=0.044), and thalamic intralaminar regions (right: t=-2.79,p=0.0067, left: t=-2.02,p=0.0046), was also correlated with greater AVH suppression.

Higher AVH severity (less AVH suppression) was correlated with increased FC strength to posterior regions extending to occipital cortex, including bilateral inferior occipital gyrus (right: t=2.83,p=0.0059, left: 2.09,p=0.0047) and middle occipital gyrus (t=2.17,p=0.03), and parietal cortex involving the left precuneus (t=2.21,p=0.03). Stronger FC to mesial temporal regions as amygdala (right: t=2.80,p=0.0064, left: t=2.56,p=0.012), hippocampus (right: t=3.04,p=0.003, left: t=2.98,p=0.0038), and left parahippocampal gyrus (t=2.07,p=0.041) were also correlated with higher AVH scores.

## Discussion

In this study, we have identified potential surgical DBS targets within the SNpr that are associated with more or fewer AVH when acutely stimulated during initial programming, suggesting a spatial gradient of anti-hallucination effect in and around the SNpr. Though limited by a focus on only short-term and self-reported responses that may not represent long-term outcomes, these exploratory findings can help to shape the iterative approach to development of ideal targeting for AVH suppression in schizophrenia since the degree of AVH suppression is associated with distinctive stimulation sites within the left and right SNpr and with connectivity to key thalamic subnuclei and prefrontal cortical regions.

The SNpr is one of the major output nuclei of basal ganglia forming the cortico-basal-ganglia-thalamic loop ^15^. It receives major inhibitory projections from the striatum, primarily from the caudate ^16^. The SNpr consists of a large number of GABAergic neurons sending inhibitory projections to brainstem regions, dorsal midbrain regions such as inferior and superior colliculi, and thalamus ^17^, including medial-dorsal thalamus (MD).

Changes in MD metabolic activities and volumes have been previously observed in patients with schizophrenia ^18,19^. In addition to SNpr input, MD serves an association hub receiving various sensory input from other brainstem nuclei and other limbic and basal ganglia structures ^20^. MD also sends efferent connections to prefrontal regions as such orbitofrontal cortex and other cortical regions extending to premotor, insular, and cingulate cortex ^21^. Because MD thus serves as a gateway for subcortical-cortical connections involved in limbic and associative processing, it follows that changes in thalamo-cortical connectivity have been observed in psychosis and schizophrenia ^22–26^.

These exploratory findings may suggest that SNpr-DBS may exert its effect on AVH by alteration of nigrothalamic signaling that normalizes dysfunctional thalamocortical connectivity. Our connectivity results support this as lower AVH burden correlated with increased functional connectivity with not only medial dorsal nucleus, but also its cortical projection targets such as OFC, ACC, and dorsolateral prefrontal cortex. Based on previous fMRI mapping studies in AVH, robust evidence supports the activation of language processing and higher-order cognitive areas like inferior frontal gyrus (including auditory cortex), anterior cingulate cortex, and temporal lobe during hallucinations ^27^. Our finding that structural connectivity between the SNpr VTA and the medial thalamus and its connected regions is associated with fewer AVH supports a mechanism of AVH reduction through normalization of distributed networks of subcortical and cortical regions involved in language processing. While studies reviewing connectivity directly from SNpr are lacking, the caudate, part of the dorsal striatum selectively connected to the SNpr, showed increased functional connectivity to regions as insula and inferior frontal gyrus after clozapine administration in schizophrenia ^28,29^.

A relatively lack of structural or functional connectivity does not immediately result in discounting mechanism of efficacy due to polysynaptic and parallel connectivity. Brain regions associated with acute AVH suppression where more functionally connected to the SNpr VTA stimulation region overlapped with structurally connected regions but also diverged. For example, inferior frontal gyrus and ventromedial prefrontal cortex showed increased functional connectivity with AVH suppression but no structural connectivity. This may be because FC can represent polysynaptic connectivity whereas SC typically looks at monosynaptic connectivity between regions. Our findings from the FC analysis align with prior research showing that networks involved in AVH include neural substrates not only in auditory perception, but syntax and semantical processing regions involved in language and memory such as hippocampus and amygdala ^30^. In a PET imaging study, patients experiencing AVH were reported to have increased activity in regions such as the broca’s area and hippocampus and parahippocampus ^31^. Similarly, based on a lesion mapping study reviewing functional networks, auditory hallucination was associated with increased connection to the posterior hippocampal subiculum ^32^. Abnormal engagement of hippocampus and its hyperactivity via associated circuitry also seem to play a role in other psychosis beyond AVH from abnormal contextualizing of sensory information ^33,34^. From our study, increased connectivity to these “psychosis” network hubs involving hippocampal and amygdala regions were associated with relatively less suppression and higher AVH burden. This raises the question of whether SNr-DBS acts by modifying networks associated with AVH generation, or instead, by engagement of compensatory networks involving DLPFC and other prefrontal regions that allow “top-down” suppression of AVH.

AVH is driven by dysfunction in distributed network across cortical regions involved in language processing and auditory perception that are likely lateralized in function. There is evidence that increased auditory hallucination severity has been associated with decreased language lateralization ^35–37^, such that there is relative hyperactivation of right hemisphere and hypoactivation of the left hemisphere. Complementary to this pattern, our structural connectivity and fiberfiltering results illustrate a strong lateralized effect such that streamlines from left SNpr stimulation were associated with more AVH suppression, and right stimulation with less AVH suppression. Because group-level differences in AVH scores between entire left and right-side stimulation were not significant **(Figure1C)**, it is reasonable that engagement of selective streamlines passing through SNpr may drive AVH depending on the hemisphere of stimulation. In AVH and schizophrenia, lateralization of structures such as supramarginal gyrus ^38^ and angular gyrus ^39,40^ has been demonstrated. Similarly, we found that acute AVH-suppression was associated with increased connectivity between the stimulation VTA and the left angular gyrus and right supramarginal gyrus. It may be possible that SNpr-DBS rebalances the weakened lateralization of circuits involved in language processing.

Another indication of the lateralized effects of SNpr DBS is that AVH suppression was maximal with stimulation in the left anterior dorsolateral SNpr, which was the opposite for the right SNpr where posterior-ventral stimulation was most effective. This inversely of “sweetspot’ and “sourspot” location between right and left SNpr may be attributed to the lateralization of aforementioned circuitry involved in language processing. In addition to the lateralized circuitry across hemisphere, the gradient of clinical outcomes may also be attributed to the topography of ipsilateral connections made from SNpr. Investigations on topographic connections within human SNpr are sparse, but early studies on non-primate models have illustrated overlapping populations of nigrotectal, nigroreticular, and nigrothalamic occupying distinctive zones within SNpr ^41,42^. Following the anterior-posterior axis, nigrothalamic cell projecting to ventroanterior-ventrolateral (VA-VL) and MD thalamus diminish in population. Following the medial-lateral axis within the first anterior part of the SNpr, nigrotectal cells projecting to colliculi regions heavily populate the lateral margin of SNpr. Based on this topography, one may theorize that our “sweetspot” for left SNpr DBS may be at a region with intersecting neuronal populations forming nigrothalamic and nigrotectal connections. Given the significance of these two circuitries in forming the cortico-thalamic-cortical loop, future radiographic studies are strongly encouraged to validate whether stimulating these two pathways achieve sufficient AVH benefits with a larger sample size and with chronic outcomes.

Several limitations exist in this study. First, our analysis was performed based on unvalidated Likert-style clinical ratings from three patients. When performing DBS programming, quick assessments are often needed to test each setting in a timeline the patient can tolerate. Validated psychosis scales cannot be repeated for each of the stimulation parameters tried during the duration of visit that most patients can tolerate. Given the experimental nature of our study with a novel surgical target for AVH in schizophrenia, a restricted number of highly refractory patients were eligible to undergo the procedure and with such a small sample size our results may have limited generalizability. However, through postoperative monopolar review montage, we were able to test numerous stimulation settings and resulting AVH scores. By incorporating each AVH scores paired with stimulating settings as individual datapoints, and pooling these scores together from all three patients, we could maintain the power of our analysis. We also tested for potential inter-patient bias of AVH scores by validating that difference in AVH scores across three patients were non-significant and by accounting for intra-patient correlation of AVH scores as a covariate in our analyses.

Furthermore, due to the nature of monopolar review, the clinical outcomes recorded were based on acute stimulation effect, and may not accurately reflect outcomes after chronic DBS, since the collection of longitudinal data on these subjects is ongoing. We will attempt to validate and update our current findings after we have sufficient datapoints during chronic stimulation in a subsequent study. Another major limitation is that we used structural and functional connectomic data generated from aggregated dMRI and fMRI sequence from group of healthy controls. Schizophrenia and AVH is considered a distributed network disorder, reflected by changes in whole-brain white matter properties derived from dMRI ^43–45^ or BOLD activation patterns ^46,47^. Using a normative connectome may thus not capture disease-specific alterations of connectivity patterns and limit the generalizability of our findings. However, similar to other network-mapping studies in movement disorders ^48,49^ and neuropsychiatric disorders ^50,51^ that utilized normative connectomes, our goal was to explore neural network substrates implicated in AVH and DBS. Nevertheless, future investigations should study SNpr DBS connectivity associated with AVH suppression using patient-specific fMRI or dMRI data.

In summary, we have identified stimulation volumes in and around the SNpr that are associated with acute AVH suppression. Many of the prefrontal and language processing regions identified in the structural and functional connectivity analyses herein are also implicated in imaging studies of AVH, and our work supports a causal relationship between AVH and these stimulation related regions. Our study may inform future iterative efforts to establish an optimal target in or near the SNpr that maximally suppresses AVH.

## Methods

### Patient Recruitment

We included three patients with TR-SZ and chronic AVH who had received investigational bilateral electrode implantation (Medtronic 3387 leads) targeting the SNpr (Clinical Trial: #NCT02361554). The patients had a diagnosis of SZ as determined by a review of medical records, discussion with referring psychiatrist as well as the Structured Clinical Interview for DSM-V (SCID-V). They were determined to be treatment-resistant for at least one year prior to the screening visit as demonstrated by clinical evidence of persistent auditory hallucinations and/or delusions that had not responded to treatment with three adequate regimens of antipsychotic medication including one failed trial of clozapine. Adequate trials of two different antipsychotic drugs belonging to different classes of at least 12 weeks equivalent to at least 500 mg/day of chlorpromazine within the previous 5 years and a trial of clozapine for at least 12 weeks at a dose of at least 400 mg (or a clozapine level of at least 350 ng/mL) were required. Patients who were unable to tolerate clozapine at this dose or for this duration because of intolerable side effects were also eligible. Patients had at least a score of 6 (severe) on 2 of the 4 Brief Psychiatric Rating Scale or BPRS positive symptoms (conceptual disorganization, grandiosity, hallucinatory behavior and unusual thought content).

One month after electrode implantation each patient underwent a systematic monopolar review stimulating each electrode contact level at escalating amplitudes while reporting their hallucination severity scores based on the Likert Scale (0 – 10). Pulse width and frequency were kept constant at 60 μsec and 130 Hz, respectively. To allow for group-wise analysis hallucination scores reported for each patient were transformed into z-scores.

### Electrode and Volume of Tissue Activation Modeling

Bilateral electrodes from three patients were localized and modeled using the Lead-DBS computational pipeline ^52^. Postoperative CT sequences were co-registered with preoperative MRI sequences using Statistical Parametric Mapping (SPM). These images were normalized into a template ICBM MNI152 brain space using Automated Normalization Tools (ANTs). Based on the metal artifact shown in the CT image, a 3D electrode model was rendered.

Based on stimulation conditions surveyed during the monopolar review for each patient an electric field model was simulated based on Finite-Element-Modeling (FEM)-based algorithm using StimFit / FieldTrip ^53^. Binary volumes of tissue activation (VTAs) were generated by thresholding regions where the magnitude of the electric field was above 0.2 V/mm ^54–56^.

### Mean Effect Image

In order to characterize how stimulation locations and stimulation volumes in the SNpr target contribute to acute AVH score during monopolar review, Z-scored AVH scores were mapped on each associated VTA and spatially averaged across all patients. This resulted in “Mean Effect Image (MEI)” for the left and right SNpr, where each voxel corresponds to the expected average AVH score after stimulation. To avoid type I error, only voxels occupied by more than 25% of total number of VTAs on each stimulation side were included in the MEI. To precisely localize regions associated with maximum and minimum acute AVH suppression, centroid coordinates representing the top and bottom 5% voxels of MEI were quantified and visualized. Centroid coordinate with top 5% voxels were considered as “sourspot” and bottom 5% voxels as “sweetspot”. All voxel-wise coordinates reported following the standard MNI coordinate system.

### Structural Connectivity Analysis

To explore how structural connections from the stimulated SNpr target are associated with AVH score, each VTA was also seeded into a normative structural connectome composed of 1,600,000 streamlines produced from diffusion MRI of 32 healthy controls as part of the Human Connectome Project (HCP)^57^. After seeding VTAs into the connectome, we generated whole-brain “fiber” streamlines and a structural connectivity (SC) map where each voxel corresponds to a number of fibers passing through the VTAs. Similar to the FC analysis, for each voxel, a multiple linear regression model was fitted by regressing SC across all patients against their respective z-scored AVH scores with patient ID as grouping variables. The resulting t-score of the beta-coefficients occupying each voxel of the brain was used to produce a “T-map” analogous to the FC approach.

### Fiberfiltering Analysis

Similar to structural connectivity analysis, fiberfiltering analysis can be used to estimate how each radiographic streamline “fiber” relative to VTAs is associated with clinical benefits of DBS ^50,58^. For each individual streamline from the normative structural connectome (identical to the one used for the structural connectivity analysis) we tested whether targeting that fiber was more likely to result in AVH score changes. VTAs corresponding to the monopolar review stimulation settings were used to classified whether or not each fiber was stimulated or not by that VTA. Across these two groups – “stimulated” and “unstimulated” – their respective AVH z-scores were compared using a two-sample t-test, resulting in a t-value assigned to each fiber. The magnitude and direction of the t-value indicates the normalized degree of AVH based on whether that fiber is stimulated or not by a VTA, such that a positive t-score is associated with relatively higher AVH score indicating less acute DBS efficacy, while a negative t-score is associated with relatively lower AVH score and more DBS efficacy. Only fibers connected to more than 10% but less than 90% of total number of VTAs were considered to reduce potential type II and type I statistical errors.

For each fiber, after performing a two-sample t-test between AVH z-scores in unconnected and connected VTAs, only fibers assigned to the top 20% and the bottom 20% of t-scores were considered for subsequent analysis. To account for potential laterality effect across hemisphere, identical methods were applied separately for left-sided VTAs and right-sided VTAs.

### Functional Connectivity Analysis

Seed-based functional connectivity was explored by seeding VTAs into a normative fMRI connectome. Whole-brain functional connectivity (FC) maps were derived by calculating voxel-wise correlation coefficients (r) of BOLD signals relative to the VTA. The normative fMRI connectome was created from fMRI sequences of 1,000 healthy adults derived as part of the Brain Genomics Superstruct Project (GSP-1000)^59^.

For each voxel, a multivariate linear regression model was fitted by regressing functional connectivity (FC) strength (r) across all patients against their respective z-scored AVH scores. In this model, patient ID (e.g., 1, 2, 3) corresponding to each VTA was included as a categorial grouping variable to account for potential inter-patient confounding effects between FC and resulting AVH changes. The resulting beta coefficient from the model fit between FC and AVH scores was transformed into a t-score for each voxel. This process was repeated for all voxels to create a “T-map,” where voxels with t-scores greater than 0 indicate regions where an increase in functional connectivity is associated with more AVH (i.e., less suppression), and voxels with t-scores less than 0 indicate regions where an increase in functional connectivity is associated with a less AVH (i.e., more suppression).

## Data Availability

The data that support the findings of this study are available from the corresponding author upon reasonable request.

## Code Availability

Codes that were written and utilized for analysis in this study are available from the corresponding author upon reasonable request.

## Acknowledgements

K.A.M. received research funding from the National Institutes of Health (NIH) /National Institute of Neurological Disorders and Stroke (NINDS; 5K23NS101096-01A1), Michael J. Fox Foundation, Parkinson’s Foundation, UCB Pharma, and Food and Drug Administration (FDA; U01FD005942) and received honoraria from the Parkinson Study Group. M.J.K received research funding from National Institutes of Health (NIH) / University of Pennsylvania School of Medicine as part of T32 Medical Scientist Training Program (MSTP).

## Author Contributions

M.J.K. conceived, organized, and executed the research project, designed and executed the statistical analysis, and wrote the first draft of the manuscript. A.B. conceived and organized the research project, and reviewed and critiqued the statistical analysis and critiqued the manuscript. Y.S. reviewed and critiqued the manuscript. A.S. reviewed and critiqued the manuscript. M.F. reviewed and critiqued the statistical analysis and critiqued the manuscript. K.S.C reviewed and critiqued the statistical analysis and critiqued the manuscript. D.S. conceived and organized the research project. K.A.M. reviewed and critiqued the statistical analysis and critiqued the manuscript. N.C. conceived and organized the research project, and reviewed and critiqued the statistical analysis and critiqued the manuscript.

