## Supplementary Material 1 for "Effective Stimulation Sites and Networks for Substantia Nigra Pars Reticulata Deep Brain Stimulation for Auditory Verbal Hallucinations in Schizophrenia"

| **Patient ID** | **Sex** | **Primary Diagnosis** | **Neuropsychiatric**  **Comorbidities** | **Handedness** |
| --- | --- | --- | --- | --- |
| **1** | Female | Schizophrenia  (Paranoid Type) | Obsessive-Compulsive Disorder (OCD)  Psychogenic Nonepileptic Seizure | Right |
| **2** | Female | Schizophrenia (Undifferentiated Type) | Obsessive-Compulsive Disorder (OCD) Posttraumatic Stress Disorder (PTSD) | Right |
| **3** | Male | Schizophrenia (Undifferentiated Type) | None | Left |

**Supplementary Material 1. Demographic and Clinical Information**
